# Evaluating the Outcomes of Enhanced Adherence Counselling Intervention on Clients with High Viral Loads in Selected Health Facilities in Monze District

**DOI:** 10.1101/2021.12.18.21267844

**Authors:** M. Kaira, T. Sikazwe, J. Simwanza, M. Zambwe, P. J Chipimo

**Author notes:** **<u>CORRESPONDING AUTHOR DETAILS</u>** Name: Mowa Zambwe, Address: Workers Compensation Fund Control Board, Lusaka, Zambia, Email ID, Guarantor of Submission: The corresponding author is the guarantor of submission.

## Abstract

To investigate the changes in Viral Load(VL) during Enhanced Adherence Counselling (EAC) sessions and its determinants among ART clients with unsuppressed VLs in Monze district.

**Method:** A Cross-sectional study involving 616 HVL ART clients from 15 health facilities in Monze district which was conducted between October 1 2019 and March 30 2021.

**Results:** Out of 616 clients analysed, there was an improvement in viral load suppression following completion of EAC with a final outcome of 61% suppression. 28.7% remained unsuppressed. A total of 9.1% had no final viral load results documented and 0.2 % had been transferred out of their respective facilities and were not included in the study. Collection of repeat Viral loads was done on 84% of the clients with high viral load results while 16% had no record of sample collection. A total of 56 results were not received giving a result return of 89% from repeat samples collected. Females had a 40% likelihood of being unsuppressed at 95% CI (41% to 86%) compared to the males.

**Conclusion:** EAC improves the outcomes of HVLs and should be encouraged on all high viral clients. Programs should be developed to improve suppression in females on ART

## INTRODUCTION

In 2014, UNAIDs released the 90,90, 90 ambitious targets towards HIV epidemic control. The first 90 being that 90% of all people living with HIV must know their status, 90% of them must be on antiretroviral therapy (ART) and 90% of them must be virally suppressed. As indicated in a study in sub-Saharan Africa 2017, almost 22 million of 36·9 million people living with HIV globally have successfully initiated antiretroviral therapy (ART).^1^ For the individual and public health benefits of ART to be realised, antiretroviral programmes, previously focussed on ART initiation, must retain patients in care and achieve high rates of viral load (VL) suppression. This requires optimizing management of those failing ART. The last further stipulates that 90% of those on ART treatment should have a suppressed VL in order to eliminate HIV transmission and prevent morbidity and mortality.^2^ To be HIV virally suppressed means to have less than 1000 copies of viruses in the body. Other than that an individual is defined to have an elevated or high viral load.^3^ According to UNAIDs, there is no definite answer to how long a person needs to take Antiretroviral (ARVs) medicines before they are virally suppressed. Regular viral load testing is therefore, necessary to ensure proper treatment monitoring on the success of the regimen. Routine viral load (VL) monitoring is the most important tool for assessing a patient’s response to treatment, and assessing adherence to antiretroviral therapy.^4^

The World Health Organization (WHO) recommends a VL test six and twelve months after starting ART and every twelve months thereafter. According to the Zambian national ART guidelines 2020, an unsuppressed Viral load result (HVL) which is as a client on ART whose viral load is above 1000 copies must undergo enhanced adherence counselling to identify virological barriers. Since the introduction of the 90, 90, 90 strategy there has been a general reduction in new HIV infections. According to UNAIDs HIV new infections as of 2019 for all ages stood at approximately 51000 and 6000 amongst children 0-14 years old from 60000 and 10000 in 2010 respectively. Despite the decline in new positive HIV infections, there is still need to prevent further infection of HIV. One of the ways in which this can be done is through the U=U strategy. The U=U (Undetectable=Untransmissible) campaign was launched as a result of four large studies conducted from 2007 to 2016 which involved thousands of serodiscordant couples that did not show a single case of sexual HIV transmission from a virally suppressed partner. The idea that someone living with HIV, who is both on treatment and virally suppressed, cannot transmit the virus to a sexual partner is revolutionary and guided decision making. According to WHO consolidated guidelines in 2013, U=U messaging had the potential to reduce stigma toward people living with HIV (PLHIV), including self-stigma; increase demand for HIV testing and antiretroviral therapy (ART), including early initiation of treatment; and improve adherence. The concept of U=U could also strengthen advocacy efforts for universal access to effective treatment and care, and messaging around U=U as a result was well-integrated into HIV prevention, care, and treatment programs, including those serving key populations.

The Zambian ART consolidated guidelines states that a person with an unsuppressed viral load is put through Enhanced adherence counselling (EAC).^4^ WHO defines treatment adherence as “the extent to which a person’s behaviour – taking medications, following a diet and/or executing lifestyle changes – corresponds with agreed recommendations from a health care provider for ART and it can therefore be said that high level of sustained adherence is necessary to suppress viral replication and improve immunological and clinical outcomes; decrease the risk of developing ARV drug resistance; and reduce the risk of transmitting HIV. Enhanced adherence counselling (EAC) is a program that offers counselling of high viral load clients on ART treatment before concluding on the effectiveness of the selected ART regimen. It is stated again in the Zambian ART guidelines that a person with a VL of 1000copies or greater is given a 1-month dispensation of ART medication while undergoing EAC sessions, a post Viral load test is then performed after 3 months to ascertain whether adherence was the cause of treatment failure^5^. However, there are various challenges associated with full completion of EAC sessions, including no documented repeated VL test result and no proper conclusion on the status of the clients.

In countries that are resource-limited such as Zambia whose HIV prevalence among women and men age 15-49 as of 2018 Zambian demographic and health survey, 2018 was at 11.1% (CI: 10.3%-11.9%). The Zambian population based HIV impact assessment showed that among People living with HIV ages 15 to 59 years who self-report current use of ART, 89.2 percent are virally suppressed: 89.7 percent of HIV-positive females and 88.2 percent of HIV-positive males who self-report current use of ART are virally suppressed.^5^ The gap in the viral load suppression till represents a population that is actively transmitting the HIV virus and it is for that reason that monitoring the change in viral load count among clients is useful for both clinicians and the nation at large as EAC reduces the chances of being switched to a second line ART regimen which is more costly. The information is needed to decide whether a significant viral load suppression before switching to a second-line regimen. A study similar to ours in Swaziland reviewed that there is a lack of evidence on the sociodemographic and clinical determinants of change in viral load count after EAC sessions among patients who enrolled in the EAC program (PLHIV with high viral load count).^2^ Therefore, it is important to evaluate the change in VL count during EAC sessions and its determinants among PLHIV with unsuppressed VL count.

The District health information system reports shows that Monze district has one of the highest people on ART a total of 18330 as of march.^6^ According to the MER report, in the past six months, the district reported 723 new HIV positives from the health facilities. This is an alarming number which shows that there is still a continuous spread of infection despite the prevention measures being undertaken. This is why the study aims at assessing the outcome of EAC sessions on ART clients with high viral load in monze district, so as to identify gaps and provide recommendations in the management of these clients.

## METHODS

### Study design and settings

This was a cross-sectional study that was used to review clients with high viral loads from health information systems and facility high viral load registers. The process of data collection involved reviewing the high viral load registers from 15 high volume facilities in Monze for clients presenting with high viral load from a period of 1^st^ October 2019 to 30^th^ March,2021. The information in these registers was then compiled and computed on an excel spreadsheet for data analysis.

### Study participants and sample size

Participants were selected from among persons with a viral load of 1000 copies or greater and were accessing treatment from a particular health facility with a current on treatment of 170 clients or greater. This was from a period of 1^st^ October 2019 to 30^th^ March 2021. A cluster sampling method was used, the clusters being geographic areas then simple random sampling was done in these clusters. This was done in order to get a sample that would be representative of the whole district. The 15 Health facilities in the district were used to form the clusters used in the study.

A sample size of 616 ART was used in the study calculated using the Slovin’s formula at 95% confidence interval, with 5% margin of error. With an assumption of 15,995 clients currently on treatment from the high-volume sites and a 96% viral suppression according to the MER and DATIM reports. Sample size calculation (Slovin’s Formula)

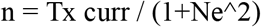

where

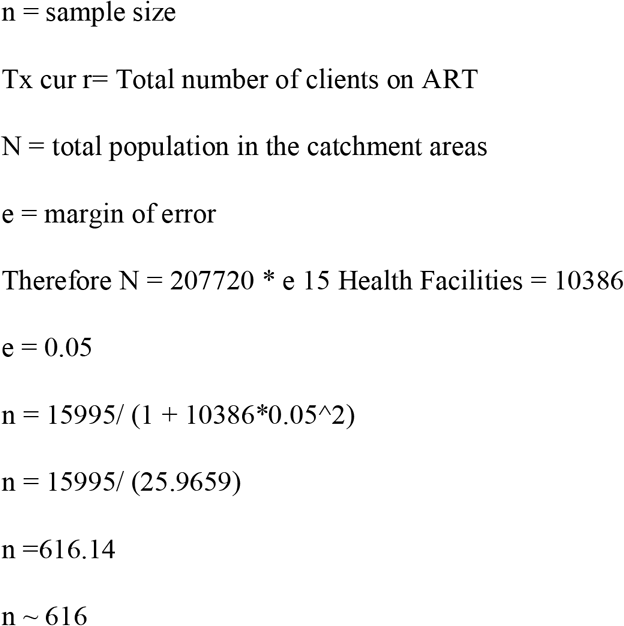

### Patient and public involvement

All ART clients were de-identified as they were entered in excel spread sheets. Consent was sought from the district health director and all health facility in-charges as data collection progressed. Approval from the ethics committee was sought from the school of medicine and health sciences study ethics committee reference; IORG00010092, University of Lusaka from 28^th^ May 2021 to 30^th^ September, 2021

### Data analysis

Analysis of continuous variables was done using frequency tables, proportions and the median was used to describe the data as it was not normally distributed. Multivariant analysis was used to deal with confounders. The shapiro-wilk test and histograms were used to check for the normality of the continuous variables. Bivariate analysis and multivariate logistic regressions were done in R. Only the variables that were significantly associated with outcome during the bivariate analysis were included in the final multivariate logistic regression. The outcome variable was suppressed and unsuppressed dichotomized as suppressed and unsuppressed. The independent variable was sex, age and each EAC session was analysed separately.

## Results

### Demographical data

The data consisted of clients of clients both male and female across all age groups with a minimun age of 5 years old and the oldest age being 77years old. Out of the 616 clients analyzed, the mean age for unsuppression was 29.5, median age was 29. 57% (n=315) of the clients were female whereas 43% (n=265) were male. 28% of the the clients were below 15 years old (n=145) whereas 78% (n=441).

### Comparison of suppressed and unsuppressed viral load before and after EAC

**Figure 1:**
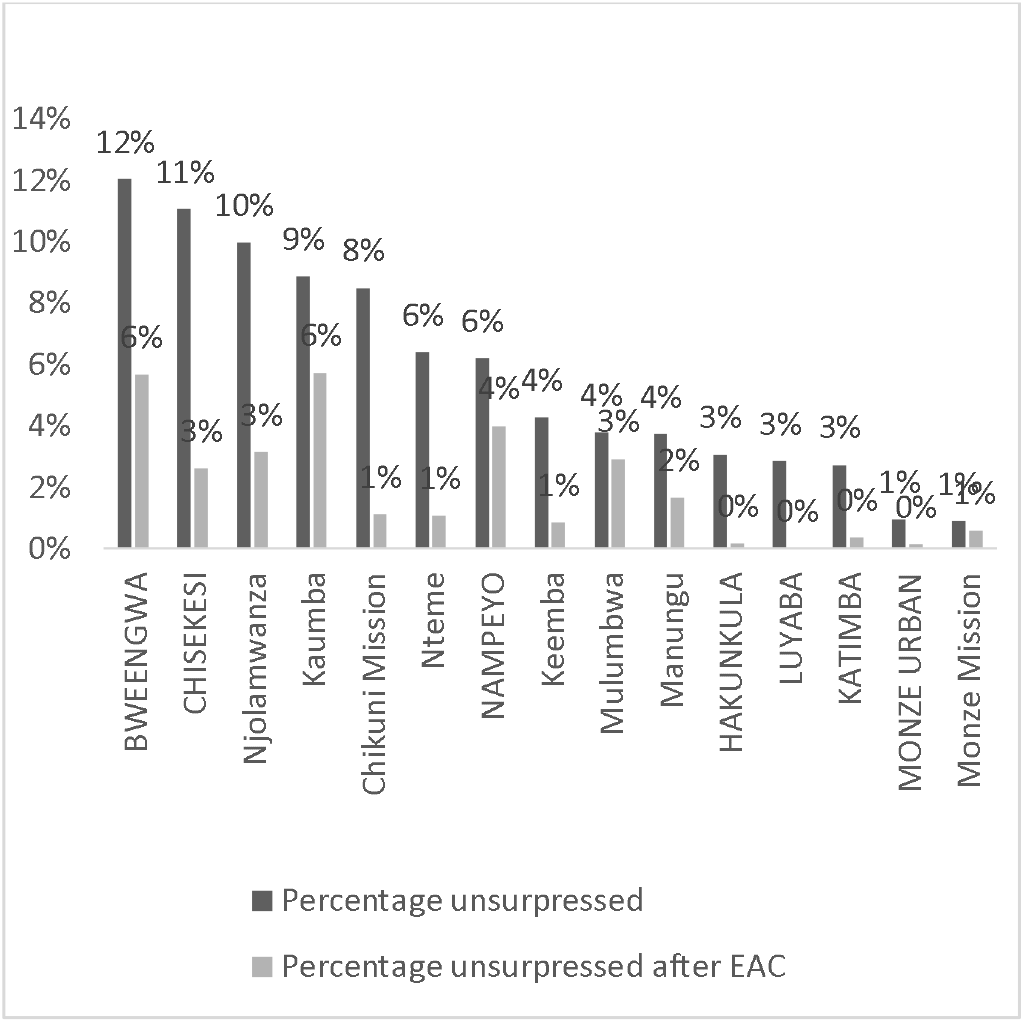
Percentage of unsuppressed clients before and after EAC. The graph shows a comparison in the suppression levels of ART clients per facility, showing a general reduction in the percentage of unsuppressed following EAC. Facilities such as Luyaba, Katimba, Hakunkula has a completely reduced their unsuppression rate to 0% following completion of EAC.

**Figure 2:**
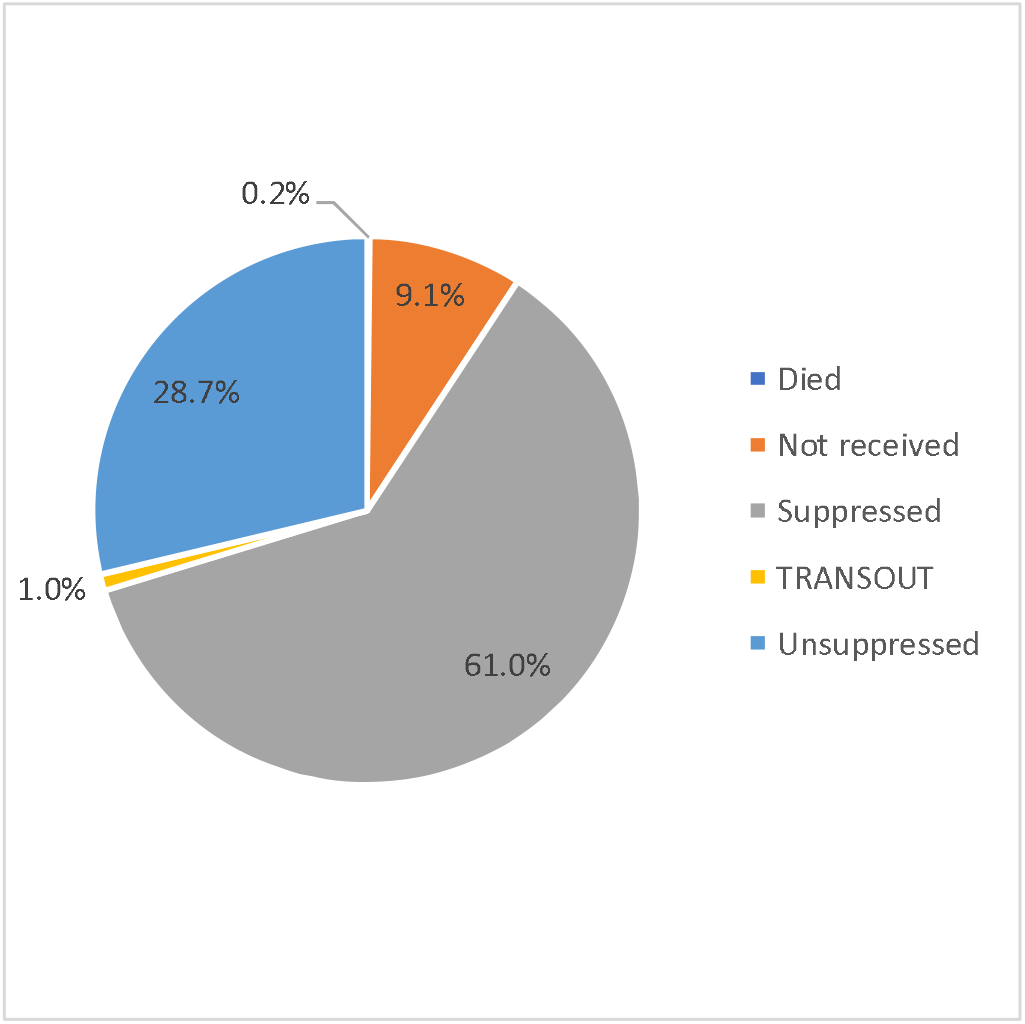
Pie chart showing outcomes after EAC. The pie chart above shows the outcomes of enhance adherence counselling with 61% of clients showing suppression after EAC. A similar study by Gedefaw et al (2020) showed a 66.4% suppression following EAC(60-72.4).

**Table 1:**
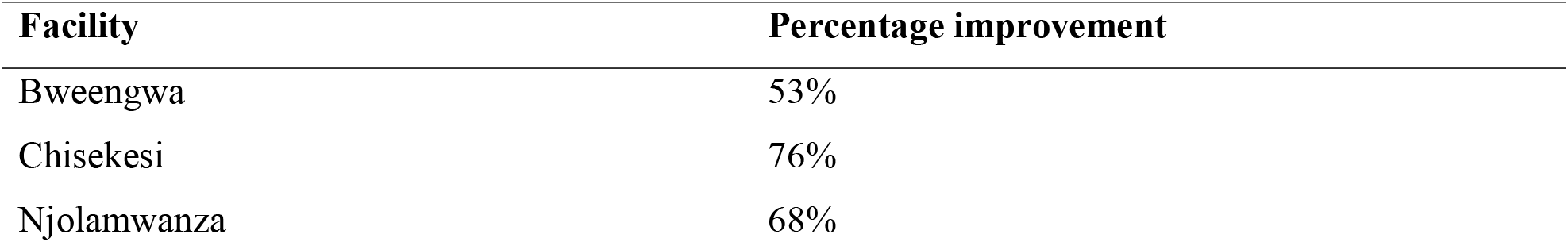
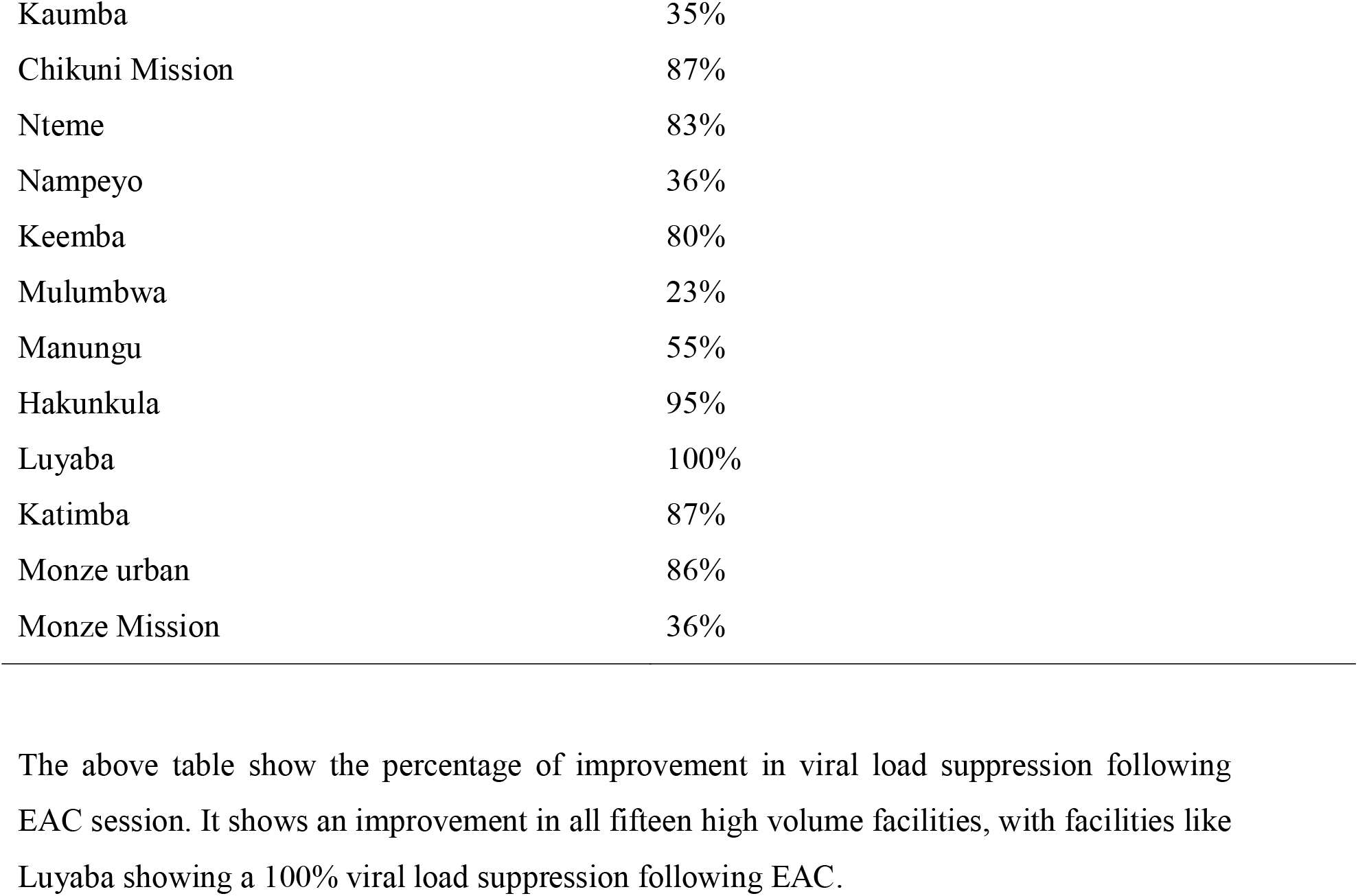
Percentage improvement of viral load Coverage following EAC in high volume facilities in Monze October 2019-30^th^ March 2021.

| Facility | Percentage improvement |
| --- | --- |
| Bweengwa | 53% |
| Chisekesi | 76% |
| Njoramwanza | 68% |
| Kaumba | 35% |
| Chikuni Mission | 87% |
| Nteme | 83% |
| Nampeyo | 36% |
| Keemba | 80% |
| Mulumbwa | 23% |
| Manungu | 55% |
| Hakunkula | 95% |
| Luyaba | 100% |
| Katimba | 87% |
| Monze urban | 86% |
| Monze Mission | 36% |

The above table show the percentage of improvement in viral load suppression following EAC session. It shows an improvement in all fifteen high volume facilities, with facilities like Luyaba showing a 100% viral load suppression following EAC.

**Table 2:** Action for the unsuppressed by facility.

| **Facility** | **Died** | **None** | **Switched to second**<br>**line** | **Total** |
| --- | --- | --- | --- | --- |
| Bweengwa | 2 | 29 | 0 | 31 |
| Chikuni Mission | 2 | 12 | 9 | 23 |
| Chisekesi | 2 | 4 | 6 | 12 |
| Hakunkula | 0 | 1 | 0 | 1 |
| Katimba | 0 | 2 | 0 | 2 |
| Kaumba | 0 | 11 | 0 | 11 |
| Keemba | 0 | 7 | 0 | 7 |
| Manungu | 1 | 25 | 15 | 41 |
| Monze Mission | 0 | 6 | 7 | 13 |
| Monze urban | 0 | 3 | 0 | 3 |
| Mulumbwa | 0 | 10 | 0 | 10 |
| Nampeyo | 1 | 8 | 0 | 9 |
| Njoramwanza | 0 | 12 | 0 | 12 |
| Nteme | 0 | 2 | 0 | 2 |

### Challenges Associated with Provision EAC

**Figure 3:**
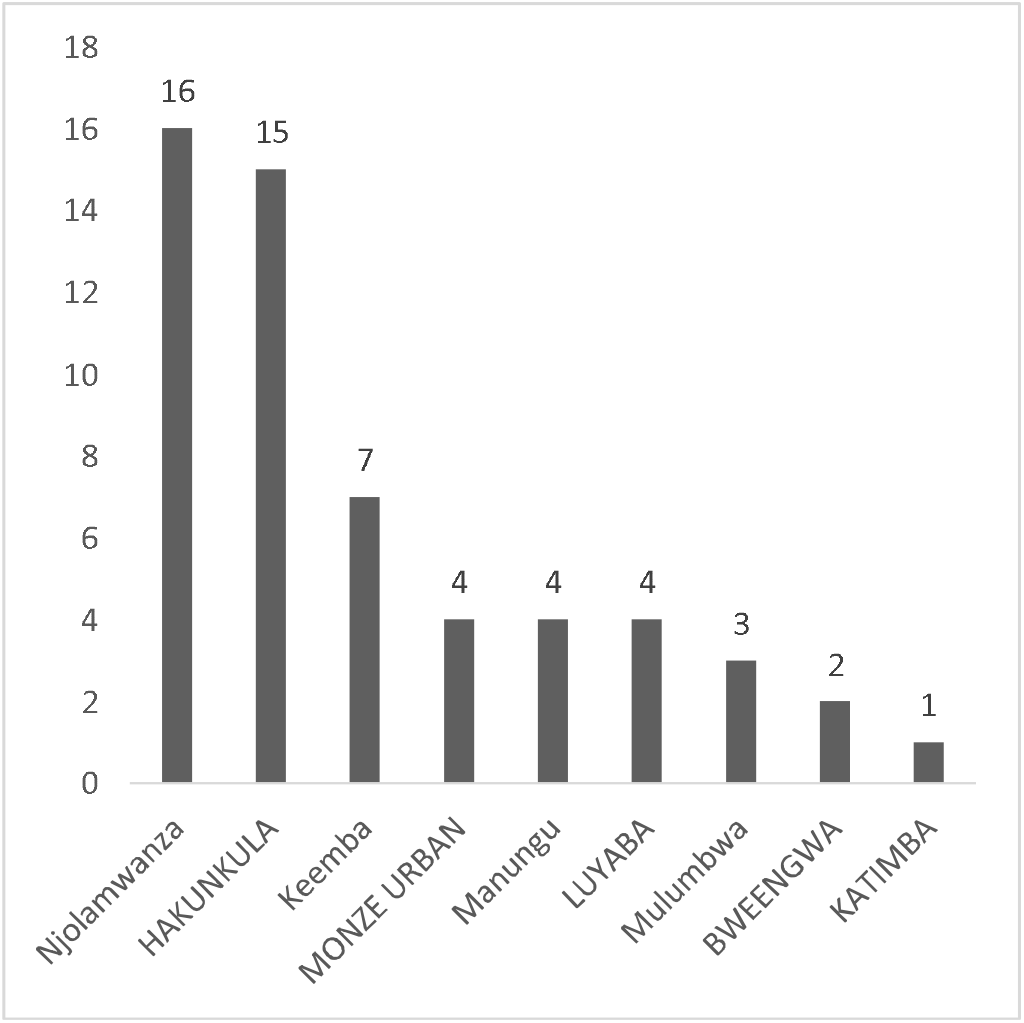
Final VL results not received per facility. The bar graph above shows the gap in management of high viral loads by showing the results not received per facility.

**Table 3:** Missed EAC sessions by facility.

| Facilities | Second EAC | Third EAC | Final EAC |
| --- | --- | --- | --- |
| Bweengwa | 4 | 6 | 10 |
| Chikuni Mission | 0 | 0 | 1 |
| Chisekesi | 0 | 0 | 0 |
| Hakunkula | 0 | 19 | 9 |
| Katimba | 0 | 0 | 0 |
| Kaumba | 0 | 0 | 0 |
| Keemba | 3 | 3 | 11 |
| Luyaba | 0 | 1 | 2 |
| Manungu | 0 | 0 | 4 |
| Monze Mission | 0 | 0 | 0 |
| Monze Urban | 0 | 0 | 0 |
| Mulumbwa | 8 | 9 | 9 |
| Nampeyo | 0 | 0 | 0 |
| Njoramwanza | 0 | 0 | 0 |
| Nteme | 0 | 0 | 2 |

The table above shows missed EAC sessions per facility. With a high missed appointments been seen by Hakunkula rural health centre.

### Logistic regression output

**Table 4:** Multivariate Logistic regression model output.

| Predictors | Outcome |  |  |
| --- | --- | --- | --- |
|  | Odds Ratios | CI | P |
| Sex |  |  |  |
| Male | Reference | Reference | Reference |
| Female | 0.6 | 0.41 – 0.86 | 0.007 |
| Second EAC |  |  |  |
| No | Reference | Reference | Reference |
| Yes | 1.43 | 0.27 – 7.88 | 0.67 |
| Third EAC |  |  |  |
| No | Reference | Reference | Reference |
| Yes | 1.63 | 0.44 – 5.76 | 0.448 |
| Final EAC |  |  |  |
| No | Reference | Reference | Reference |
| Yes | 2.15 | 0.84 – 5.46 | 0.103 |

The females had 40% increased likelihood of being unsuppressed at 95%CI (41% to 86%) compared to the males. Though conducting EAC sessions showed reduced likelihood of being unsuppressed at 95%CI (Second EAC 70%, Third EAC 61% and Final EAC 47%), this was not statistically significant.

## DISCUSSION

In this study, out of the 616 clients analyzed, the mean age for un-suppression was 29.5, median age was 29. 57% (n=315) of the clients were female whereas 43% (n=265) were male. 28% of the the clients were below 15 years old (n=145) whereas 78% (n=441). Out of the 616 clients analyzed, the mean age for unsuppression was 29.5, median age was 29. 57% (n=315) of the clients were female whereas 43% (n=265) were male. 28% of the the clients were below 15 years old (n=145) whereas 78% (n=441).

And out of 616 clients analysed, there was an improvement in viral load suppression following completion of EAC with a final outcome of 61% documented as suppressed, 28.7% remained unsuppressed, 9.1% had no final viral load results documented and 0.2 percent had been transferred out of their respective facilities. A similar study in Nigeria showed that among two thirds (71.9%) of clients with high viral loads undergoing EAC only 51% were suppressed.^7^ Which is similar to cases in Zimbabwe and South Africa. Another study in sub-saharan Africa showed 8.1% increase in suppression following EAC (95% CI: 0.1% to 17.2%)^8^. Even though our findings were not statistically significant, which could be attributed to the fact that our study faced challenges arising from incomplete data and only complete and well documented EAC sessions were included in the logistic regression. These studies also reviewed the challenge of no collection of repeat viral loads post three months of EAC sessions, a finding that is similar to that of our findings.^8, 9^

Females had a 40% likelihood of being unsuppressed at 95% CI(41% to 86%) compared to the males. The finding is consistent with a study in Mozambique that had a 61.2% likelihood of females to be unsuppressed.^10^ The finding could also be debated on as the majority of clients on treatment are females as compared to male, hence the need to open up more differentiated service delivery models that will encourage male clinical attendance. The median age for un suppression was 29.6 years, which was similar with a finding in study that showed with 36.6% of those unsuppressed being paediatrics while 60.3% being adults (above 15years of age).^9^

Out of the 616 clients with High viral loads analysed, only 23% of them were documented to have been switched to second line. This could have been attributed to both a gap in knowledge of management of high viral loads or a gap in supply chain management causing stock out of the second line regimen. Only four high volume sites recorded switching to second line regimen and these were Chisekesi, Chikuni, Manungu and Monze mission hospital which gives us the key facilities to focus on and mentor based on this gap. The study attributed it to low viral load testing and suggested that a change in strategy or a differentiated care based on whether a person is virologically suppressed is required is need to meet the targets. A quality improvement collaboration project in South Africa showed in retrospective data showed that only 22% patient had unsuppressed VL received 3 documented EAC sessions and only 35% of them had a repeat viral load contrary to our study had had 84% recollection of repeat viral load for those unsuppressed. This study however, further showed that hospitals achieved better switching rates than health facility which is some in common with our study, with about 90% switching rate to second line.^10^

Collection of repeat Viral loads was done on 84% of the clients with high viral load results while 16% had no record of sample collection. Challenges with no repeat viral load testing mainly arise from poor documentation or lack of feedback of final results. A total of 56 results were not received giving a result return of 89% from repeat high viral load samples collected. This possesses a challenge for health care workers to finally conclude on the outcomes of high viral loads and further cannot decline whether to conclude treatment failure or not. There is therefore need to strengthen laboratory feedback systems so as to improve patient management.

Although conducting EAC sessions showed reduced likelihood of being unsuppressed at 95%CI (Second EAC 70%, third EAC 61% and final EAC 47%), this was not statistically significant. This was not consistent with other studies, such as a study in South Africa that showed that out of a total of 165 patients that were enrolled in adherence counselling following suppression (81.8% female, median age 36.2 years) and at the closure of the study, 119 patients (72.0%) were virally suppressed and 148 patients (89.0%) were retained in care. Six, 12 and 18 months after adherence counselling enrolment, retention in care was estimated at 98.0%, 95.0% and 89.0%, respectively.^11^ Viral suppression was estimated to be maintained by 90.0%, 84.0% and 75.0% of patients at 6, 12 and 18 months after adherence counselling enrolment, respectively. Showing significantly those patients who struggled to achieve or maintain viral suppression in routine clinic care can have good retention and viral suppression outcomes in adherence counselling, a differentiated ART delivery model, following suppression support. Another study in Kenya showed that clients with high viral loads 75.7% enrolled in EAC and 31.2% received VL suppression.^12^ There are major programmatic gaps being faced in receiving full completion of EAC, a finding that was similar to our study. A study in Uganda attributed the poor performance showed only 23% of the clients undergoing intensive adherence counselling achieved suppression, this they attributed to poor counselling skills from health care providers.^13^ The shortfall of our study however was lack on quality data to assess the effectiveness of Enhanced adherence counselling which could be a direction for future studies. Additional barriers could have been from incomplete data in registers which were the primary data collection tools in this study.

## CONCLUSION AND LIMITATIONS

### Conclusion

We can conclude that there was an improvement in the outcomes of high viral load clients undergoing enhanced adherence counselling. Though this was not statistically significant under a 95% CI and this can be attributed to the various limitations in the study which included: incomplete post Viral load result documented in the register, incomplete EAC sessions and poor documentation of the client’s final outcome. There is therefore need for the district health team to provide technical support to health facilities so as to improve the outcomes of Enhances adherence counselling especially in resource limited countries such as Zambia were the costs of second line treatment is high, re-suppression as a result of EAC sessions will be useful.

### Limitations

The limitations to this study were the incomplete filling in of high viral load registers in most health facilities in Monze which led to limitations in analysis of some data elements such as the outcomes of EAC. Another limitation was missing data on variables such as age, ART start date that limited the analysis of other variables that could have made the study broader. There was also a challenge of missing high viral load registers at facilities such Monze and Manungu urban clinics and this prevented us from using the high viral load registers in certain periods in which the timeframe corresponded to the prescribed period in the study methodology, this could have led to the leaving out significant of data elements that may have not affected our study outcomes. The study also did not include any competency assessments for the health care workers providing EAC, this could have also affected the outcomes and can be added to future studies.

## Supporting information

strobe check list

## Data Availability

All data produced in the present study are available upon reasonable request to the authors

## Ethical considerations

Approval was obtained from the University of Lusaka. Additionally, approval was obtained from the National Health Research Authority. All participants in the study were required to participate voluntarily by providing informed consent, they were assured that no harm would come upon them should they decide not to take part in the study and they were not required to provide their personal identification information.

## AUTHOR CONTRIBUTIONS

MK

- Work conception. Data acquisition, analysis and interpretation.
- Manuscript drafting.
- Approval of final manuscript.
- Accountable for all aspects of the work regarding its accuracy or integrity.

TS

- Work conception, Data analysis and interpretation.
- Manuscript drafting.
- Approval of final manuscript.
- Accountable for all aspects of the work regarding its accuracy or integrity.

MZ

- Manuscript drafting.
- Critical revisions for intellectual content.
- Data interpretation
- Approval of final manuscript.
- Accountable for all aspects of the work regarding its accuracy or integrity.

JS

- Manuscript drafting.
- Critical revisions for intellectual content.
- Data interpretation
- Approval of final manuscript.
- Accountable for all aspects of the work regarding its accuracy or integrity.

PJC

- Manuscript drafting.
- Critical revisions for intellectual content.
- Data interpretation
- Approval of final manuscript.
- Accountable for all aspects of the work regarding its accuracy or integrity.

## References

1. Shroufi, A. Van Custem, G. Cambiano, V. Bansi-Matharu, L. Duncan, K. Murphy, R Maman, D. Phillips, A. (2019), Simplifying switch to second line ART: Predicted effect of defining failure of first-line efavirenz-based regimens in sub-Saharan Africa by a single viral load > 1000 copies/ml. Copyright ©Wolters Kluwer Health, Inc.

2. Gedefaw, D. Melese, L. (2020), Change in viral load count and its predictors among unsupressed VL patients receiving EAC intervention at three hospitals in northern ethopia; An exploratory retrospective follow up study, HIV/AIDs study and palliative care 2020:12 869–877

3. Etoori, D. Ciglenecki, I. Ndlangamandla, M. Edwards, CG. Jobanputra, K. Pasipamire M. (2018), Successes and challenges in optimizing the viral load cascade to improve antiretroviral therapy adherence and rationalize second-line switches in Swaziland. J Int AIDS Soc; 21(10):1–8

4. Adait, T. (2016), Global AIDS update, Geneva: UNAIDS; (http://www.unaids.org/en/resources/documents/2016/Global-AIDS-update-2016, accessed

5. Zambian Guidelines for treatment and prevention of HIV infection (2020), Ministry of health-Zambia, p50-62 9MOH-Zambia).

6. Population-Based HIV Impact Assessment (2016) ’Zambia Population-Based HIV Impact Assessment’, Journal of the International AIDS Society, (December 2016), pp. 1–4. Available at: http://phia.icap.columbia.edu/wp-content/uploads/2016/09/ZAMBIA-Factsheet.FIN_.pdf. 7 https://spho.more-zm.org:8450/dhis-web-dataentry/index.action

7. Olutosin, A. Awolude, Oluwatobi, O. Moradeyo, M. Abiolu, J. (2021), Virologic Outcomes Following Enhanced Adherence Counselling among Treatment Experienced HIV Positive Patients at University College Hospital, Ibadan, Nigeria, STD Study & Reviews, 0(1): 53–65, 2021; Article no. ISRR.65271ISSN: Pages 2347–5196, NLM ID: 101666147

8. El-Sadr WM et al (2017). Realizing the potential of routine viral load testing in sub-Saharan Africa Journal of the International AIDS Society, 20(S7):e25010

9. Finci1, A. Flores, A. G. Gutierrez Zamudio1, A. Matsinhe1, E. de Abreu1, S. Issufo, Gaspar, I. Cigleneck L. Molfino (2020), Outcomes of patients on second-and third-line ART enrolled in ART adherence clubs in Maputo, Mozambique, Tropical Medicine and International Health, volume 25.

10. Joseph S. (2019), Outcomes of patients enrolled in an antiretroviral adherence club with recent viral suppression after experiencing elevated viral loads, published Online:https://doi.org/10.4102/sajhivmed.v20i1.905

11. Miriam, R. Dunstan, A. Steve, A. Rodrigo, B. Maureen, K. Isaac, L. Caitlin, M-M. Redempta, M. Lilly, N. Onyango C. (2020), Improving Utilization of HIV Viral Load Test Results Using a Quality Improvement Collaborative in Western Kenya, September-October • Volume 31 • Number 5 Journal of the Association of Nurses in AIDS Care

12. Nasuuna et al (2018), Low HIV viral suppression rates following the intensive adherence counseling (IAC) program for children and adolescents with viral failure in public health facilities in Uganda, BMC Public Health 18:1048 https://doi.org/10.1186/s12889-018-5964-

